# Quantitative susceptibility mapping for detection of kidney stones, hemorrhage differentiation and cyst classification in ADPKD

**DOI:** 10.1101/2024.02.24.24303322

**Authors:** Karl Schumacher, Martin R. Prince, Jon D. Blumenfeld, Hanna Rennert, Zhongxiu Hu, Hreedi Dev, Yi Wang, Alexey V. Dimov

**Author notes:** Corresponding author: Alexey V. Dimov, PhD MRIRI, 407 E 61^st^, Suite RR107, New York, NY, 10021, USA.

## Abstract

**Background and purpose:** The objective is to demonstrate feasibility of quantitative susceptibility mapping (QSM) in autosomal dominant polycystic kidney disease (ADPKD) patients and to compare imaging findings with traditional T1/T2w magnetic resonance imaging (MRI).

**Methods:** Thirty-three consecutive patients (11 male, 22 female) diagnosed with ADPKD were initially selected. QSM images were reconstructed from the multiecho gradient echo data and compared to co-registered T2w, T1w and CT images. Complex cysts were identified and classified into distinct subclasses based on their imaging features. Prevalence of each subclass was estimated.

**Results:** QSM visualized two renal calcifications measuring 9 and 10 mm, and three pelvic phleboliths measuring 2 mm but missed 24 calcifications measuring 1mm or less and 1 larger calcification at the edge of the field of view.

A total of 121 complex T1 hyperintense/T2 hypointense renal cysts were detected. 52 (43%) cysts appeared hyperintense on QSM consistent with hemorrhage; 60 (49%) cysts were isointense with respect to simple cysts and normal kidney parenchyma, while the remaining 9 (7%) were hypointense. The presentation of the latter two complex cyst subtypes is likely indicative of proteinaceous composition without hemorrhage.

**Conclusions:** Our results indicate that QSM of ADPKD kidneys is possible and uniquely suited to detect large renal calculi without ionizing radiation, and able to identify properties of complex cysts unattainable with traditional approaches.

## 1. Introduction

Autosomal dominant polycystic kidney disease (ADPKD) is the most common hereditary kidney disease with progressive renal function decline affecting ∼1 in 1000 subjects within the general population [1-3] and accounting for ∼10% of patients with end-stage renal disease (ESRD) requiring chronic dialysis or renal transplantation [4]. The disease is characterized by uncontrolled formation and growth of fluid-filled cysts leading to compression and destruction of nephrons, interstitial fibrosis and subsequent renal failure. Currently, there is no cure for the disease, but tolvaptan, a Type 2 vasopressin receptor blocker, can reduce the rate of cyst growth and forestall renal failure.

The most common clinically relevant complications of ADPKD, apart from renal failure, are bleeding and nephrolithiasis [5]. Recent findings indicate that cystic hemorrhage is a marker of the ongoing kidney injury[6] and is indicative of rapid progression of chronic kidney disease (CKD)[6-8]. While conventional magnetic resonance imaging (MRI) is sensitive to the presence of hemorrhage, it is highly nonspecific due to the similar appearance of benign proteinaceous cysts [6, 9, 10]. MRI is also ill-suited for detection of kidney calcifications, which can be associated with significant pain and morbidity[11] and indicative of higher risk of ESRD[12, 13]. Non-contrast CT has high sensitivity and specificity for renal calcifications[14], but has low specificity for identification of hemorrhage[15], and is not appropriate for periodic screening due to cumulative effects of ionizing radiation[16].

Quantitative susceptibility mapping (QSM) [17, 18] identifies local tissue susceptibility sources such as calcium (a negative susceptibility source) or hemorrhage (a positive susceptibility source). Susceptibility maps are extracted from multi-echo gradient echo (GRE) images through spatial deconvolution according to the dipole field model in magnetism physics [19, 20]. QSM differentiates calcifications from hemorrhages and protein deposition on the basis of their magnetic properties [18, 21-23]. Despite the major potential benefits of QSM in assessment of ADPKD pathology, incorporating QSM into clinical abdominal imaging remains a challenge due to time constraints, respiratory motion, chemical shift of fat and strong magnetic field inhomogeneity within the abdomen and pelvis [22]. The aim of the present study is to demonstrate the feasibility of QSM in ADPKD patients and to compare imaging presentations of tissue pathologies on QSM and traditional T1- and T2-weighted MRI. To address the technical challenges of abdominal QSM, we extracted QSM from the breath hold multiecho chemical shift encoded GRE sequence that is routinely acquired in all abdominal MRI patients for characterizing fat, water and iron deposition thereby eliminating the need to acquire any additional data. To overcome the challenge of magnetic field mapping in the presence of chemical shift we utilized the MR sequence echo time asymmetry (some echo more in-phase and some echos more out-of-phase) to perform iterative separation of water and fat signals [24, 25] with graph cuts unwrapping initialization to create pure water local field maps [26] to be inverted for mapping susceptibility.

## 2. Materials and Methods

### Study population

This study was performed under an Institutional Review Board (IRB) approved retrospective protocol. All subjects met the Pei criteria for ADPKD [27, 28] with at least 3 cysts on ultrasound prior to 40 years of age and with a positive family history of ADPKD or exclusion of all other causes of polycystic kidneys in the absence of family history. Raw data suitable for QSM processing was successfully retrieved from 33 of 34 consecutive ADPKD patients; in one subject with severe motion artifacts, QSM could not be performed. Demographic data on the 33 subjects included in the present study are shown in Table 1.

**Table 1.** ADPKD patient characteristics at the time of image acquisition.

|  |  |
| --- | --- |
| Age (years) | 40.3±11.8, 95%CI: [36.3...44.3] |
| Sex, male : female | 11 : 22 |
| creatinine | 1.16±0.52, 95%CI: [0.98...1.34] |
| eGFR | 76.99±31.34, 95%CI: [66.45...87.52] |
| **mutations** |  |
| PKD1 | 14 |
| PKD2 | 2 |
| IFT140 | 2 |
| unknown | 15 |
| Total Kidney Volume | 1537±1081, 95%CI: [1173...1900] |
| **Mayo Imaging Classification** |  |
| 1A | 1 |
| 1B | 6 |
| 1C | 12 |
| 1D | 11 |
| 1E | 3 |
| 2 | 0 |
| CT shows Calcifications | 7 |

### Imaging protocol

and underwent imaging at 3T on a clinical MR scanner (GE Healthcare, Waukesha, WI) using a body phased array coil between July 2021 and March 2023. The imaging protocol included axial and coronal T2-weighted fast spin echo sequences (voxel size = 0.78×0.78×4 mm^3^, echo time (TE) = 92 ms, repetition time (TR) = 1010 ms, flip angle (FA) = 130°, readout bandwidth (rBW) = 355 Hz/pixel) and a 3-dimensional multiecho gradient echo IDEAL/IQ sequence (matrix size = 224×224×30 interpolated to 512×512×60; voxel size = 0.78×0.78×4 mm^3^, first TE = 1.2 ms, echo spacing (ΔTE) = 1.1 ms, #TE = 6, TR = 8.3 ms, rBW = 390 Hz/pixel). This axial multiecho GRE acquisition was performed as two acquisitions each with a separate breathhold, to cover the entire abdomen and pelvis. In 8 subjects T1-weighted 3D FSPGR sequence acquired with the following parameters: voxel size = 0.74×0.74×3 mm^3^, TE = 1.8 ms, TR = 4.9 ms, FA = 12°, rBW = 558 Hz/pixel. All acquisitions were performed using breath-hold technique. Additionally, patient’s clinical history was reviewed for abdominal CT examinations within 10 years prior to or after the MR imaging.

### Image reconstruction

To avoid patient discomfort and an increase in examination time for additional dedicated sequences, QSM was reconstructed from the GRE data routinely acquired to generate water, fat and 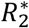 images [29] existing in the patient records. This was possible because raw k-space data is now routinely saved from the scanner for at our institusion for all IDEAL/IQ sequences. Individual coil images for each echo time were reconstructed, combined using sum-of-squares and corrected for gradient non-linearity distortions using vendor-distributed software toolbox. To estimate the frequency maps, the multiecho phase data were fitted to a nonlinear model [30]. Then the result was spatially unwrapped using graph cuts allowing no fat/water overlap [26], and used to initialize water-fat separation with simultaneous mapping of the magnetic field [24] to eliminate the contribution of fat chemical shift to the signal. To estimate tissue field distribution, background magnetic field was removed using projection onto the dipole field technique [31]. QSM was reconstructed using Morphology-Enabled Dipole Inversion (MEDI)[19]. The reconstruction pipeline (Figure 1) was implemented in Matlab (Mathworks, Natick, MA, USA). All DICOM images were converted to NIFTI format for further processing. Images were co-registered into GRE image space using ITK-SNAP[32]. When necessary due to non-rigid deformation of the tissues and differences in breath hold depth between acquisitions, registration was performed separately for the left and right kidneys.

**Figure 1.**
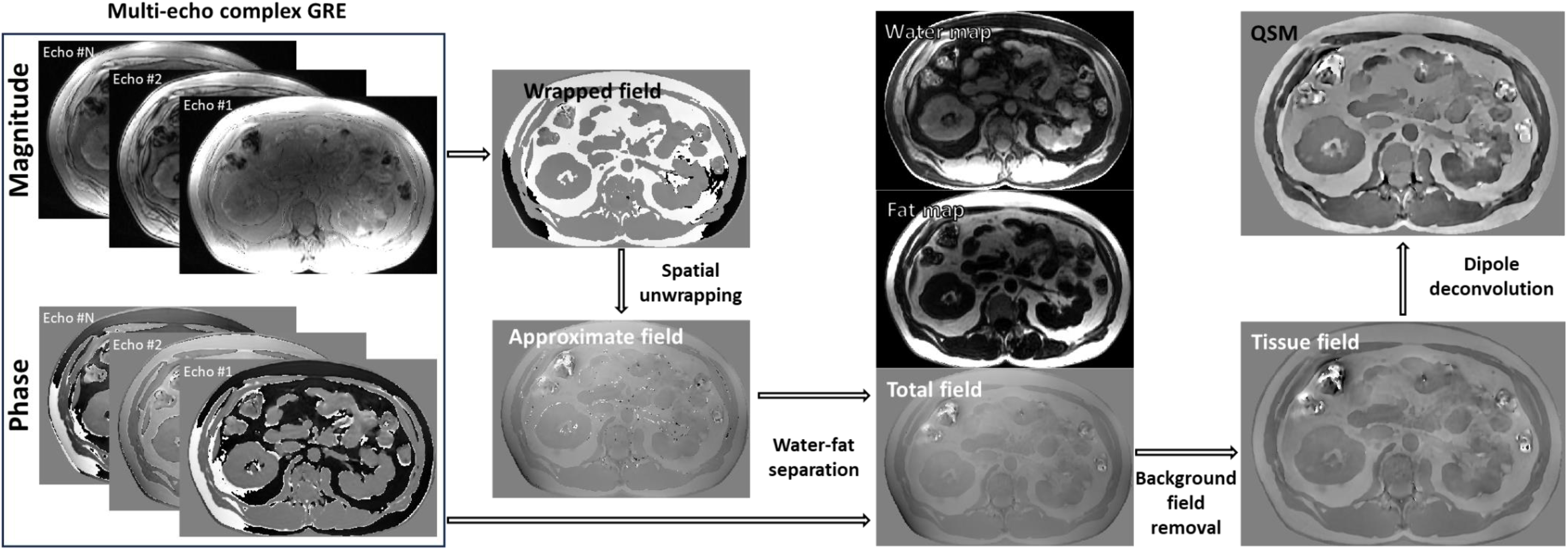
Flowchart of quantitative susceptibility mapping (QSM) reconstruction in application to ADPKD imaging. Susceptibility mapping requires acquisition and preservation of both magnitude and phase of a breathhold complex gradient echo sequence. Water-fat separation is performed to generate an accurate magnetic field map. The obtained total filed can next be decomposed into the background field and the field generated by the sources within the tissues of interest (tissue field). Dipole deconvolution to solve the inverse field-to-source problem and decode non-local field distribution into magnetic susceptibility, a local tissue property, is performed as the final step.

### Image analysis

Reconstructed susceptibility maps were reviewed in all subjects and cross-referenced against other MR contrast and CT images, whenever available. Observed lesions were assessed according to their susceptibility. Specifically, lesions with a positive value (hyperintense relative to simple fluid filled cysts and normal kidney parenchyma) on QSM images were interpreted as paramagnetic. Lesions with hypointense appearance on QSM images were interpreted as diamagnetic. To avoid misclassification of paramagnetic fat within renal sinuses as cysts in QSM, water and fat fraction maps were also used.

Subjects with simultaneously available T1w and T2w images were screened for the presence of T1 hyper- and T2 hypointense lesions consistent with the classic presentation of cystic hemorrhage [7, 33]. Identified cysts were referenced to QSM and their appearance on susceptibility maps was classified as either hypo-, iso- or hyperintense, and the number of cysts in each class was recorded.

Additionally, all QSM hyper- and hypointense cysts larger than 2 mm in diameter identified in the study cohort were counted manually, their appearance on individual T2- and T1-weighted MR images recorded, and the prevalence of the identified QSM-based cyst subclasses estimated.

### Statistical analysis

Continuous variables were described using mean and SD. Frequency and percentage were calculated for categorical variables. Trends in the QSM-visible cyst subclass populations were analyzed by determining a Pearson correlation coefficient on the incidence of each cyst subclass per patient. A Pearson correlation matrix was constructed to find the linear correlation between the incidence of each cyst subclass across all patients. The corresponding p-values were calculated to determine the correlation significance defined at the level of 0.05.

## 3. Results

### QSM vs CT findings

Abdominal CT within 10 years (median = 6 years, range: 0.5 to 10 years) of MRI was available in 12 subjects, 7 of whom had positive findings. In total, 27 calcifications were detected and comprised renal calculi (n = 15) and pelvic phleboliths (n =12). Two renal calcifications identified on CT measuring 9 and 10 mm were readily visible as hypointense on QSM (≈ −1.5 *ppm*) and dark on all MRI sequences (Figure 2A&B). One renal calcification measuring 11 mm was not visualized on QSM due to the location at the border between the two acquisition volumes, although it was discernable in the reconstructed frequency map. In one subject, CT revealed bilateral phleboliths in the pelvic area (diameter *d* ≈2mm) which were also visible on QSM. The remaining calcifications were 1 mm or less in diameter and not visible on QSM (e.g., Figure 2B, white arrows).

**Figure 2.**
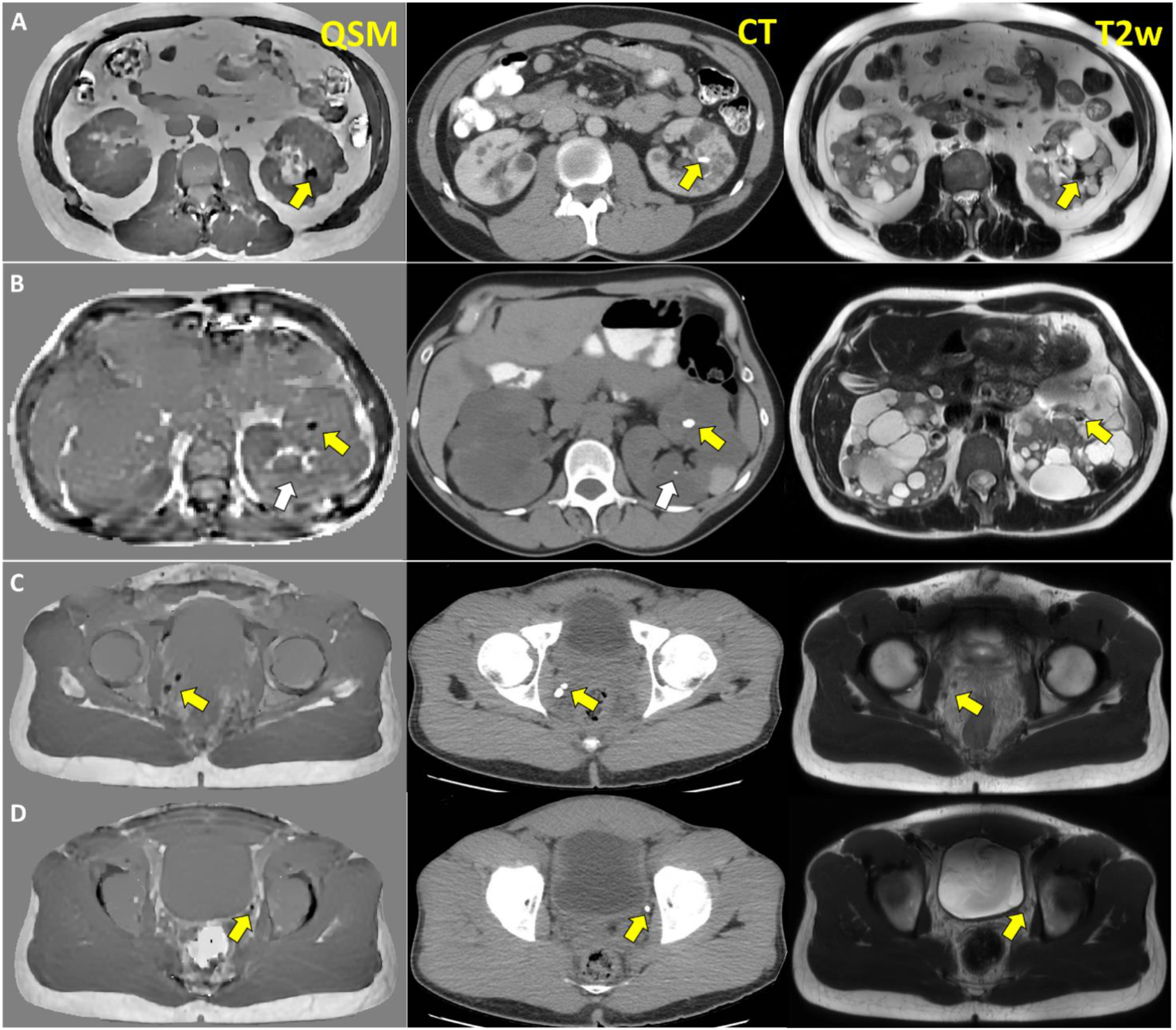
Comparison of appearance of renal calcifications (A, B) and phleboliths (C,D) on QSM, CT and T2w. On QSM calcifications appear as strongly diamagnetic regions (approximately -1.5 ppm), providing a basis for distinction between them and QSM-hypointense cysts.

### QSM in detection of hemorrhagic cysts

T1w and T2w images were simultaneously available in 8 out of 33 subjects contributing to the total of 121 T1 hyper/T2 hypointense cysts. 52 (43%) cysts appeared hyperintense (≈ 0.3 *ppm*) on QSM consistent with the presentation of hemorrhage (Figure 3A, yellow arrows); 60 (49%) cysts were isointense with respect to simple cysts and normal kidney parenchyma (Figure 3A, red arrows), while the remaining 9 (7%) were hypointense (≈ −0.2 *ppm*) (Figure 3B).

**Figure 3.**
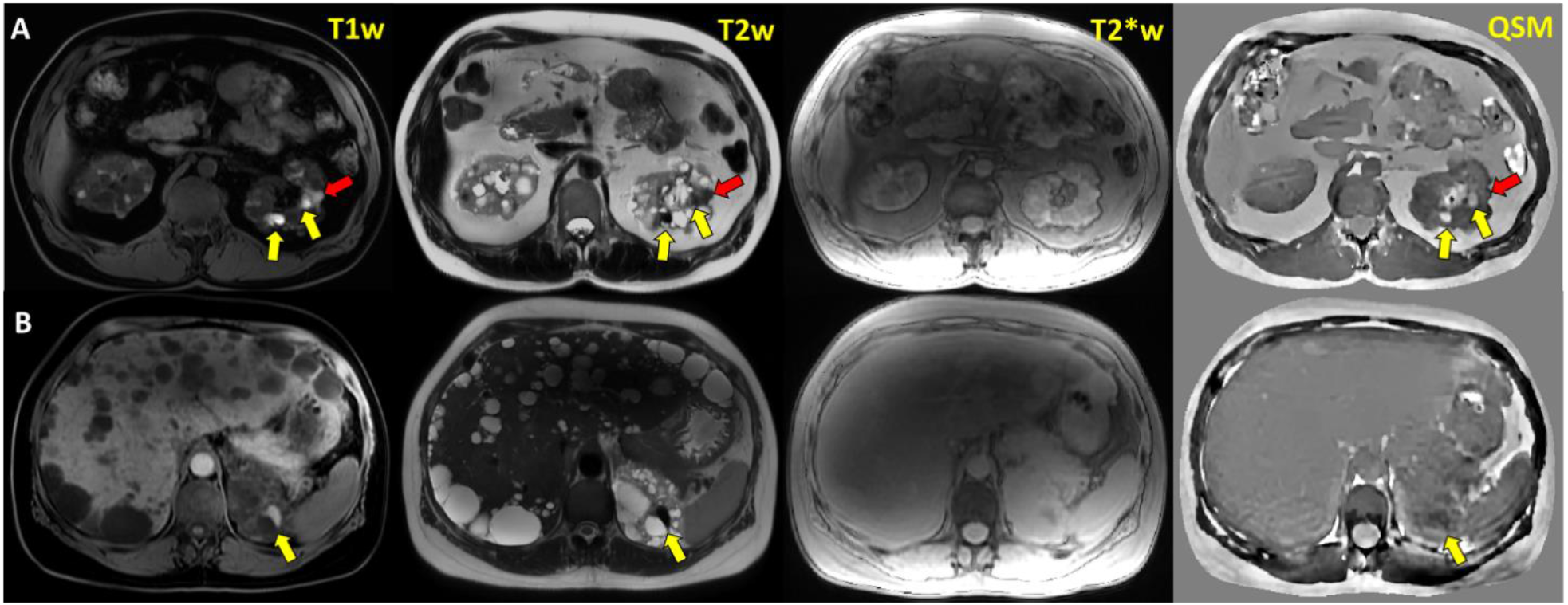
Examples of T1 hyperintense/T2 hypointense cysts with different magnetic properties as visualized by QSM. GRE-derived T2*w image naturally co-registered to QSM is provided for anatomic reference. A) Paramagnetic (≈ 0.3 *ppm*), QSM hyperintense cysts strongly suggestive of chronic hemorrhage (yellow arrows) along an isointense cyst (red arrow); B) Diamagnetic (≈ −0.2 *ppm*), QSM hypointense cyst. QSM iso- and hypointensity is indicative of non-hemorrhaging nature of T1/T2 shortening and different cysts composition.

### QSM-based cysts subclassification: QSM-visible cysts and T2w

In the complete cohort of 33 subjects, 250 QSM-visible cysts were identified. Among those, we established 6 subclasses of QSM-visible cysts based on their appearance on T2-weighted images (QSM hyper- or hypo-intense and T2 hyper-, iso-, or hypo-intense):

T2 hypointense/QSM hyperintense (“-/+”) cysts (Figure 2A) occurred with the highest incidence over the whole population with 106 catalogued cysts representing 42.4% of all QSM-visible cysts. Majority of this subclass’ population occurred in four patients (#1, 2, 6 and 5, see Figure S1), who had an incidence of 23, 17, 15, and 14 T2 hypointense/QSM hyperintense cysts. T2 hypointense/QSM hypointense (“-/-”) cyst subclass (Figure 2B) occurred with the second highest incidence of 41 cysts (16.4% of 250 of identified QSM-visible cysts). Majority of this subclass’ population occurred in two patients (#2 and 7, Figure S1) who had 5 and 7 cysts of this type. T2 isointense (with respect to kidney parenchyma)/QSM hyperintense (“=/+”) cysts subclass (Figure 3A) had the 3^rd^ highest incidence over the whole population with 38 cataloged cysts representing 15.2% of all QSM-visible cysts. The plurality of these cysts occurred in two patients (#2 and #6) who had an incidence of 6 and 7 T2 isointense/QSM hyperintense cysts, respectively. T2 hyperintense/QSM hypointense (“+/-”) (Figure 4B) cysts occurred as the subtype with the 4^th^ highest incidence (33 out of 250), representing 13.2% of QSM -complex cysts. The simple majority of this subclass population occurred in just three patients (#6, 9, and 25) who had an incidence of 5, 4, and 4 T2 hyperintense/QSM hypointense cysts. The two remaining cyst subclasses together accounting for 12.8% of QSM-visible cysts were represented by T2 hyperintense/QSM hyperintense (“+/+”, 6.4% of the total count; Figure 4C) and T2 isointense/QSM hypointense (“=/- “, 6.4% of the total count) cysts.

**Figure 4.**
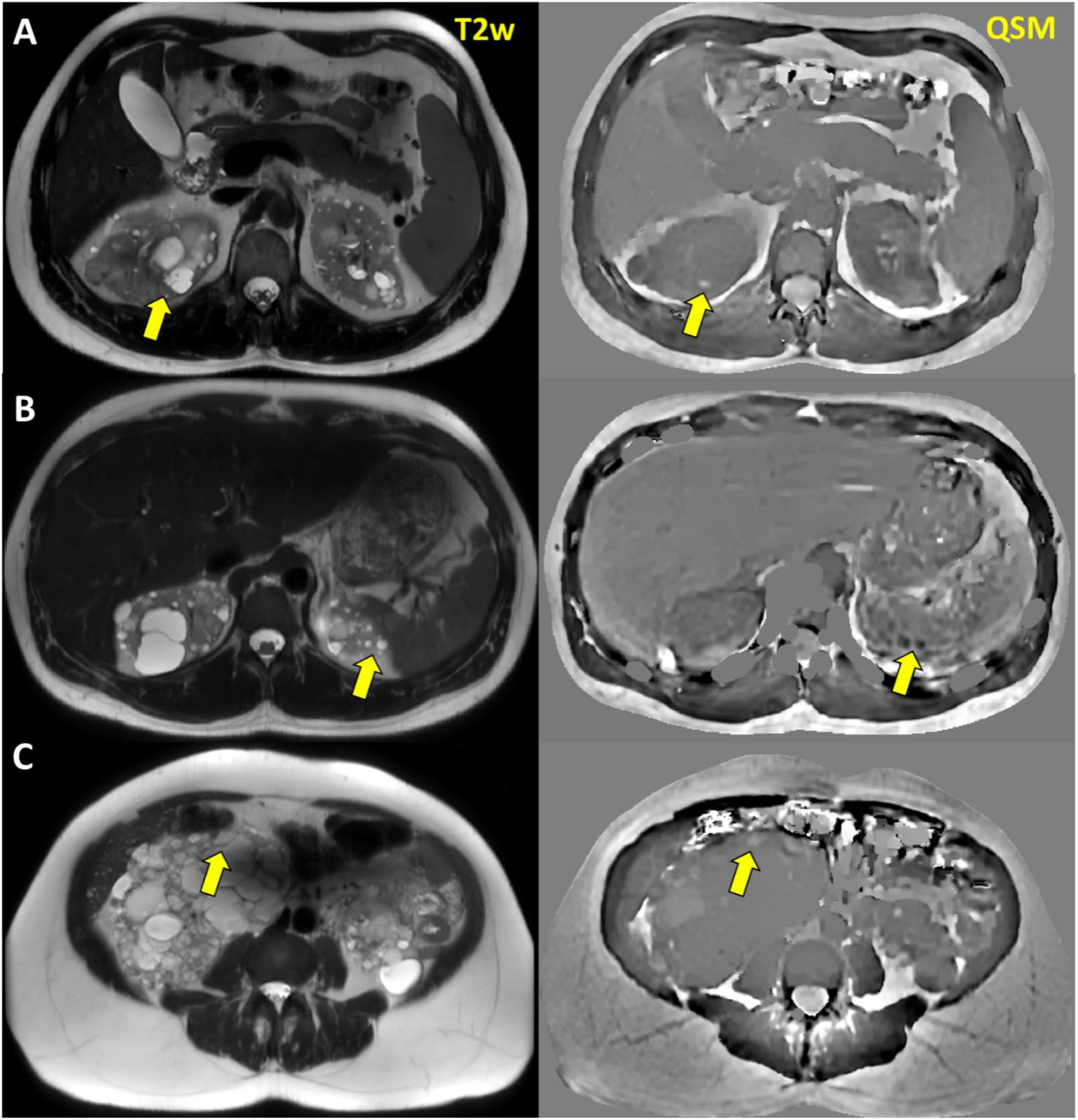
Additional examples of QSM-visible cysts with different appearance on T2w MRI. A) Paramagnetic (≈ 0.3 *ppm*) QSM hyperintense/T2 isointense cyst; B) Two diamagnetic (≈ −0.3 *ppm*) QSM hypointense/T2 hyperintense cysts; C) Weakly paramagnetic (≈ 0.09 *ppm*) QSM hyperintense/T2 hyperintense cyst.

### QSM-based cysts subclassification: QSM-visible cysts and T1w

Analysis of the subjects with available T1w images contributed 55 QSM-visible cysts. The majority of the identified QSM complex cysts occurred in subject #1 contributing 30 cysts in total. Among the identified cysts, the following 3 cyst subclasses were established:

T1 hyperintense/QSM hyperintense cysts occurred with the highest incidence rate of 60% across the 8 patients; however, when excluding the contribution of subject #1, T1 hyperintense/QSM hyperintense cysts (Figure 3A) had only 36% incidence. The T1 isointense (with respect to kidney parenchyma)/QSM hypointense (Figure 5) and T1 hyperintense/QSM hypointense (Figure 3C) cyst subclasses had the incidence rates of 12.7% and 10.9% across the 8 patients, correspondingly; excluding the contribution of patient #1, these subclasses had an identical 20% incidence rate across the 7 remaining patients.

**Figure 5.**
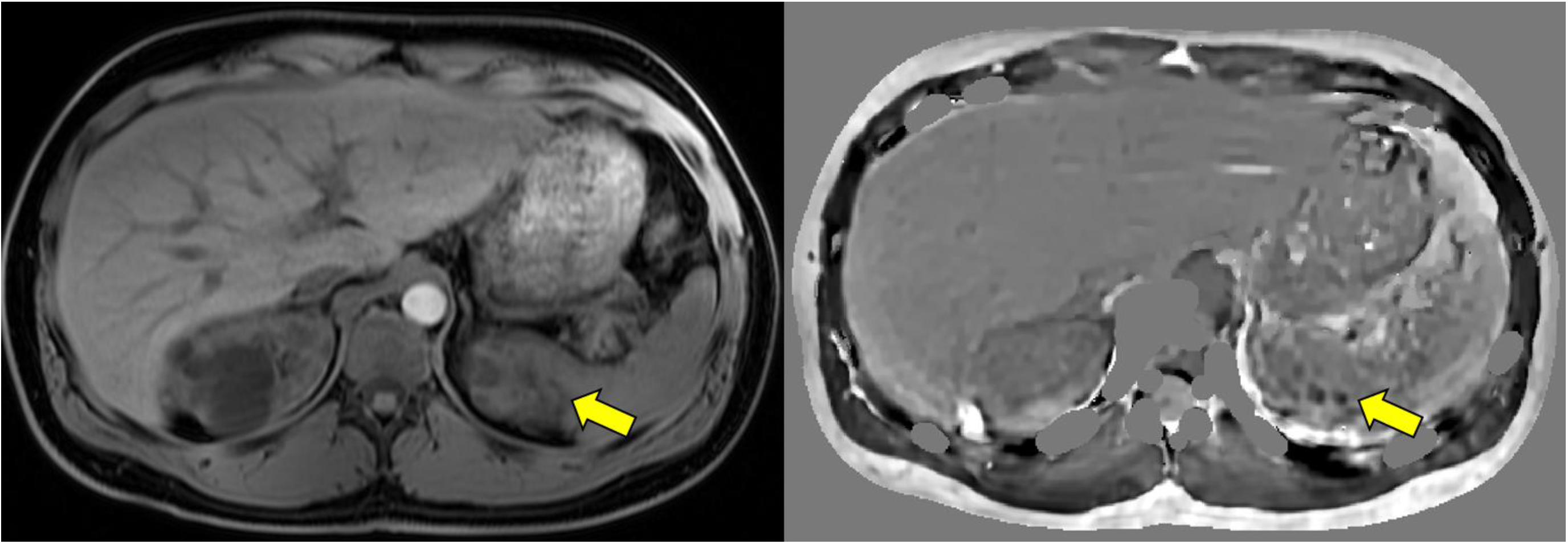
Two T1 isointense/QSM hypointense cysts

### Correlation analysis

The Pearson correlation analysis found significant positive correlations between 6 different cyst subclasses. T2 bright/QSM dark cysts and T2 bright/QSM bright cysts incidences are moderately correlated with a coefficient of 0.58 and significance p < 0.01. T2 isointense /QSM bright cysts are moderately correlated with T2 bright/QSM bright, and T2 bright/QSM dark with positive coefficients 0.48 and 0.44 respectively and both with high significance p < 0.01. T2 dark/QSM bright is moderately correlated with T2 bight/QSM bright, and T2 bight/QSM dark, and strongly correlated with T2 isointense/QSM Bright cysts with a coefficient of 0.76 and p < 0.01.

Complete results of the correlation analysis are shown in Fig. S2.

## 4. Discussion

In this pilot study, we have demonstrated the feasibility of renal QSM in 33 ADPKD subjects showing its potential to provide complimentary information to other MRI pulse sequences about cyst composition in patients with polycystic kidney disease. QSM, while well established in neurologic applications[18, 23, 34-38], remains a challenging task in body imaging. Thus, due to the common occurrence of fat-based tissues characterized by non-negligible chemical shift (≈ −3.5*ppm*, or −440 *Hz* on 3T), data acquisition for QSM requires strategic selection of acquisition parameters including echo time and readout bandwidth; nonrigid displacement of organs due to respiratory motion necessitates use of breath holding and special techniques to shorten acquisition times. In the present work we have proposed an efficient method allowing easy incorporation of QSM into abdominal MR using a widely available routine IDEAL/IQ sequence normally included in every abdominal MRI for water/fat/iron quantification. There are two major findings in our study: a) QSM can detect large clinically relevant calcifications in kidneys and the urinary tract although 1mm stones are not visualized and b) QSM provides complementary information regarding the underlying tissue pathology not accessible with either of T1w and T2w MRI, and their combination.

Over the past decades, T1w and T2w MRI have been used to non-invasively probe changes in renal microstructure and tissue composition in ADPKD[10, 39, 40]. T2-weighted MRI provides high-resolution imaging of fluid-filled renal cysts without ionizing radiation exposure. T1 and T2-weighted MRI images can differentiate between simple and complex cysts without the need for gadolinium injection [41], but are often unable to clearly discriminate between bleeding and infection or neoplasia [3, 42, 43]. The connection between voxel contents and T1/T2 relaxation times is complex and nonlinear [44, 45] and is dependent on the microscopic motion-coupling mechanisms, such as water diffusion across an inhomogeneous susceptibility fields (outer sphere effect [46]) and water proton exchange with molecules (inner sphere effect [47]). Relaxation depends on cystic proteinaceous exudates and blood products that decrease the microscopic diffusional motion of water molecules and, therefore accelerating relaxation, making T1w and T2w MRI non-specific and hard to interpret. The gradient echo (GRE) sequence without spin refocusing is sensitive to the differences in magnetic susceptibility within the underlying tissue [48, 49] and has recently seen increasing interest as a means to probe kidney parenchymal oxygenation, perfusion and fibrosis [50-53]. However, GRE data is not a direct measurement of tissue susceptibility[54]: T2*w magnitude and phase contrasts at a given voxel depend not only on the tissue susceptibility in that voxel but also on that of the nearby voxels with a distance and direction dependent weighting [55].

Presently, there are no reliable non-contrast MRI techniques for subclassification of renal cysts based on their composition. Current standard of care MR imaging is also inadequate for diagnosis of nephrolithiasis due to rapid decay of water proton signal in calculi, leading to nonspecific signal voids that are indistinguishable from air or artifacts. In contrast, MRI signal phase allows rigorous biophysical modeling for the determination of tissue magnetic susceptibility [17, 20]. Data acquisition for QSM is easily implementable on any clinical platform and does not require special hardware or sequences [17, 56].

Magnetic susceptibility as a molecular electron cloud property is linearly proportional to molecular concentration [57], permitting linear chemical decomposition for tissue characterization [58]. QSM allows for direct characterization by distinguishing the presence of blood degradation products such as hemosiderin with positive (strongly paramagnetic) susceptibility from calcifications and proteinaceous exudate with negative susceptibilities.

Indeed, this principle has been successfully demonstrated in the present work. As our data suggests, incorporation of QSM into renal imaging protocol allows detection of calcifications in kidneys. Kidney stones are common among the chronic kidney disease and ADPKD patients[59-61]. Patients with urinary stones should be followed up more closely for progression[12]. Non-contrast CT is a highly sensitive and specific technique for imaging kidney stones[14]. Once a partially obstructing ureteral stone has been identified on CT, following the gradual descent of the stone down the ureter is not practicable with CT due the cumulative radiation exposure of multiple scans[16]. QSM exploits large differences in magnetic susceptibilities of mineral depositions and that of surrounding soft tissues to distinguish diagnostic features associated with calcifications. QSM offers the potential to follow stones to see if they are progressing down the ureter on a path toward being excreted or if an intervention will be required. While some of the calcifications detected by CT were not visible on QSM even when covered by the imaging volume, all of them had diameter of 1 mm or less and not diagnostically relevant.

The second major finding of the present study is that incorporation of QSM in assessment of complex cysts might result in more specific characterization of hemorrhagic cysts. The present approach to detect cystic hemorrhage involves multiparametric MR imaging and identification of cysts characterized by T1 and T2 relaxation time shortening. However, the same shortening can result from other physical tissue properties such as the high viscosity of the proteinaceous cystic exudate. In this sense, identification of complex QSM iso- and hypointense cysts is in good agreement with previous pathology findings [15] where benign cysts containing fluid with high protein content. As proteins are diamagnetic in nature (e.g., serum albumin’s susceptibility *χ*_*SA*_=-1.3 ppm)[62], their presence can be readily distinguished from paramagnetic blood degradation products such as deoxyHb and metHb (e.g., deoxyhemoglobin’s molar susceptibility *χ*_*dHb*_=10.8 ppm)[63, 64] using QSM, but not in relaxation-based MR contrasts. Interestingly, in our sample only 43% of T1 hyperintense/T2 hypointense cysts were paramagnetic. This finding is significant in the light of recent data indicating that presence of hemorrhaging cysts is indicative of faster disease progression [7], and calls for further evaluation of role of QSM as a potential imaging biomarker in a larger patient cohort.

Our data also suggest existence of possibility for further subclassification of simple renal cysts based on their QSM appearance. Specifically, we demonstrate existence of T1- and T2 isointense para- and diamagnetic cysts (likely a result of higher sensitivity of QSM to the changes in tissue composition compared to relaxation-based contrasts [65-67]) which warrants further investigation.

A major limitation of this feasibility study is the lack of histopathological confirmation of cyst composition, limited number of CT-confirmed calcifications, and data heterogeneity resulting in relatively small number of patients with simultaneously available T1w/T2w images. In addition, comparison with QSM approaches for dealing with a large dynamic range of susceptibilities, including the STAR-QSM [68] and TFI [69] methods could not be performed. Furthermore, the present study does not incorporate magnetic source separation to disentangle contributions of para- and diamagnetic sources potentially co-existing within the cysts [70-72]. Inclusion of other clinical data such eGFR, ht-TKV and Mayo class [73] into the analysis is necessary to further elucidate the relation between the presence of QSM cyst subclasses and renal function decline. The imaging protocol used in the present study utilized thick slices that might be sub-optimal for detection of small cysts and calcifications. The two-step acquisition of the pelvic and abdominal volumes with GRE can result in loss of QSM visualization at the transition between acquisitions; a single volume centered on kidneys is preferable. Thus, further optimization of the acquisition scheme might be required. Finally, there was a considerable time interval between the CT and MRI examinations, and future work should incorporate prospective study design.

In conclusion, our study demonstrates that QSM can be utilized in ADPKD subjects to distinguish hemorrhaging and dense proteinaceous cysts, as well as to detect calcifications within the kidneys and downstream in the urinary system. Utilization of QSM in addition to the traditional multi-contrast MRI has the potential to more fully characterize the underlying ADPKD pathology. QSM can be easily incorporated into clinical imaging using standard, widely available GRE sequences used in abdominal imaging already for fat and iron quantitation.

## Data Availability

All data produced in the present study are available upon reasonable request to the authors

## Notes

### Competing Interest Statement

The authors have declared no competing interest.

### Funding Statement

This study did not receive any funding

### Author Declarations

This study was performed under an Weill Cornell Medicine Institutional Review Board's approved retrospective protocol.

## References

1. Gabow PA. Autosomal dominant polycystic kidney disease. N Engl J Med. 1993;329(5):332–42. doi: 10.1056/NEJM199307293290508.

2. Spithoven EM, Kramer A, Meijer E, Orskov B, Wanner C, Abad JM, et al. Renal replacement therapy for autosomal dominant polycystic kidney disease (ADPKD) in Europe: prevalence and survival--an analysis of data from the ERA-EDTA Registry. Nephrol Dial Transplant. 2014;29 Suppl 4(Suppl 4):iv15–25. doi: 10.1093/ndt/gfu017.

3. Torres VE, Harris PC, Pirson Y. Autosomal dominant polycystic kidney disease. Lancet. 2007;369(9569):1287–301. doi: 10.1016/S0140-6736(07)60601-1.

4. Perrone RD, Ruthazer R, Terrin NC. Survival after end-stage renal disease in autosomal dominant polycystic kidney disease: contribution of extrarenal complications to mortality. Am J Kidney Dis. 2001;38(4):777–84. doi: 10.1053/ajkd.2001.27720.

5. Cornec-Le Gall E, Alam A, Perrone RD. Autosomal dominant polycystic kidney disease. Lancet. 2019;393(10174):919–35. doi: 10.1016/S0140-6736(18)32782-X.

6. Suwabe T, Ubara Y, Sumida K, Hayami N, Hiramatsu R, Yamanouchi M, et al. Clinical features of cyst infection and hemorrhage in ADPKD: new diagnostic criteria. Clin Exp Nephrol. 2012;16(6):892–902. doi: 10.1007/s10157-012-0650-2.

7. Riyahi S, Dev H, Blumenfeld JD, Rennert H, Yin X, Attari H, et al. Hemorrhagic Cysts and Other MR Biomarkers for Predicting Renal Dysfunction Progression in Autosomal Dominant Polycystic Kidney Disease. J Magn Reson Imaging. 2021;53(2):564–76. doi: 10.1002/jmri.27360.

8. Cornec-Le Gall E, Audrezet MP, Rousseau A, Hourmant M, Renaudineau E, Charasse C, et al. The PROPKD Score: A New Algorithm to Predict Renal Survival in Autosomal Dominant Polycystic Kidney Disease. J Am Soc Nephrol. 2016;27(3):942–51. doi: 10.1681/ASN.2015010016.

9. Suwabe T, Ubara Y, Ueno T, Hayami N, Hoshino J, Imafuku A, et al. Intracystic magnetic resonance imaging in patients with autosomal dominant polycystic kidney disease: features of severe cyst infection in a case-control study. BMC Nephrol. 2016;17(1):170. doi: 10.1186/s12882-016-0381-9.

10. Marotti M, Hricak H, Fritzsche P, Crooks LE, Hedgcock MW, Tanagho EA. Complex and simple renal cysts: comparative evaluation with MR imaging. Radiology. 1987;162(3):679–84. doi: 10.1148/radiology.162.3.3809481.

11. Nishiura JL, Eloi SR, Heilberg IP. Pain determinants of pain in autosomal dominant polycystic kidney disease. J Bras Nefrol. 2013;35(3):242–3. doi: 10.5935/0101-2800.20130038.

12. Ozkok A, Akpinar TS, Tufan F, Kanitez NA, Uysal M, Guzel M, et al. Clinical characteristics and predictors of progression of chronic kidney disease in autosomal dominant polycystic kidney disease: a single center experience. Clin Exp Nephrol. 2013;17(3):345–51. doi: 10.1007/s10157-012-0706-3.

13. Alexander RT, Hemmelgarn BR, Wiebe N, Bello A, Morgan C, Samuel S, et al. Kidney stones and kidney function loss: a cohort study. BMJ. 2012;345:e5287. doi: 10.1136/bmj.e5287.

14. Brisbane W, Bailey MR, Sorensen MD. An overview of kidney stone imaging techniques. Nat Rev Urol. 2016;13(11):654–62. doi: 10.1038/nrurol.2016.154.

15. Fishman MC, Pollack HM, Arger PH, Banner MP. High protein content: another cause of CT hyperdense benign renal cyst. J Comput Assist Tomogr. 1983;7(6):1103–6.

16. . Health Effects of Exposure to Low Levels of Ionizing Radiations: Time for Reassessment? Washington (DC)1998.

17. Wang Y, Liu T. Quantitative susceptibility mapping (QSM): Decoding MRI data for a tissue magnetic biomarker. Magn Reson Med. 2015;73(1):82–101. doi: 10.1002/mrm.25358.

18. Harada T, Kudo K, Fujima N, Yoshikawa M, Ikebe Y, Sato R, et al. Quantitative Susceptibility Mapping: Basic Methods and Clinical Applications. Radiographics. 2022;42(4):1161–76. doi: 10.1148/rg.210054.

19. Liu J, Liu T, de Rochefort L, Ledoux J, Khalidov I, Chen W, et al. Morphology enabled dipole inversion for quantitative susceptibility mapping using structural consistency between the magnitude image and the susceptibility map. Neuroimage. 2012;59(3):2560–8. doi: 10.1016/j.neuroimage.2011.08.082.

20. de Rochefort L, Liu T, Kressler B, Liu J, Spincemaille P, Lebon V, et al. Quantitative susceptibility map reconstruction from MR phase data using bayesian regularization: validation and application to brain imaging. Magn Reson Med. 2010;63(1):194–206. doi: 10.1002/mrm.22187.

21. Chen W, Zhu W, Kovanlikaya I, Kovanlikaya A, Liu T, Wang S, et al. Intracranial calcifications and hemorrhages: characterization with quantitative susceptibility mapping. Radiology. 2014;270(2):496–505. doi: 10.1148/radiol.13122640.

22. Dimov AV, Li J, Nguyen TD, Roberts AG, Spincemaille P, Straub S, et al. QSM Throughout the Body. J Magn Reson Imaging. 2023;57(6):1621–40. doi: 10.1002/jmri.28624.

23. Eskreis-Winkler S, Zhang Y, Zhang J, Liu Z, Dimov A, Gupta A, Wang Y. The clinical utility of QSM: disease diagnosis, medical management, and surgical planning. NMR Biomed. 2017;30(4). doi: 10.1002/nbm.3668.

24. Fuller S, Reeder S, Shimakawa A, Yu H, Johnson J, Beaulieu C, Gold GE. Iterative decomposition of water and fat with echo asymmetry and least-squares estimation (IDEAL) fast spin-echo imaging of the ankle: initial clinical experience. AJR Am J Roentgenol. 2006;187(6):1442–7. doi: 10.2214/AJR.05.0930.

25. Reeder SB, Wen Z, Yu H, Pineda AR, Gold GE, Markl M, Pelc NJ. Multicoil Dixon chemical species separation with an iterative least-squares estimation method. Magn Reson Med. 2004;51(1):35–45. doi: 10.1002/mrm.10675.

26. Dong J, Liu T, Chen F, Zhou D, Dimov A, Raj A, et al. Simultaneous phase unwrapping and removal of chemical shift (SPURS) using graph cuts: application in quantitative susceptibility mapping. IEEE Trans Med Imaging. 2015;34(2):531–40. doi: 10.1109/TMI.2014.2361764.

27. Pei Y, Obaji J, Dupuis A, Paterson AD, Magistroni R, Dicks E, et al. Unified criteria for ultrasonographic diagnosis of ADPKD. J Am Soc Nephrol. 2009;20(1):205–12. doi: 10.1681/ASN.2008050507.

28. Ravine D, Gibson RN, Walker RG, Sheffield LJ, Kincaid-Smith P, Danks DM. Evaluation of ultrasonographic diagnostic criteria for autosomal dominant polycystic kidney disease 1. Lancet. 1994;343(8901):824–7. doi: 10.1016/s0140-6736(94)92026-5.

29. Eskreis-Winkler S, Corrias G, Monti S, Zheng J, Capanu M, Krebs S, et al. IDEAL-IQ in an oncologic population: meeting the challenge of concomitant liver fat and liver iron. Cancer Imaging. 2018;18(1):51. doi: 10.1186/s40644-018-0167-3.

30. Liu T, Wisnieff C, Lou M, Chen W, Spincemaille P, Wang Y. Nonlinear formulation of the magnetic field to source relationship for robust quantitative susceptibility mapping. Magn Reson Med. 2013;69(2):467–76. doi: 10.1002/mrm.24272.

31. Liu T, Khalidov I, de Rochefort L, Spincemaille P, Liu J, Tsiouris AJ, Wang Y. A novel background field removal method for MRI using projection onto dipole fields (PDF). NMR Biomed. 2011;24(9):1129–36. doi: 10.1002/nbm.1670.

32. Yushkevich PA, Piven J, Hazlett HC, Smith RG, Ho S, Gee JC, Gerig G. User-guided 3D active contour segmentation of anatomical structures: significantly improved efficiency and reliability. Neuroimage. 2006;31(3):1116–28. doi: 10.1016/j.neuroimage.2006.01.015.

33. Roubidoux MA. MR imaging of hemorrhage and iron deposition in the kidney. Radiographics. 1994;14(5):1033–44. doi: 10.1148/radiographics.14.5.7991812.

34. Vinayagamani S, Sheelakumari R, Sabarish S, Senthilvelan S, Ros R, Thomas B, Kesavadas C. Quantitative Susceptibility Mapping: Technical Considerations and Clinical Applications in Neuroimaging. J Magn Reson Imaging. 2021;53(1):23–37. doi: 10.1002/jmri.27058.

35. Ravanfar P, Loi SM, Syeda WT, Van Rheenen TE, Bush AI, Desmond P, et al. Systematic Review: Quantitative Susceptibility Mapping (QSM) of Brain Iron Profile in Neurodegenerative Diseases. Front Neurosci. 2021;15:618435. doi: 10.3389/fnins.2021.618435.

36. Liu C, Wei H, Gong NJ, Cronin M, Dibb R, Decker K. Quantitative Susceptibility Mapping: Contrast Mechanisms and Clinical Applications. Tomography. 2015;1(1):3–17. doi: 10.18383/j.tom.2015.00136.

37. Nikparast F, Ganji Z, Zare H. Early differentiation of neurodegenerative diseases using the novel QSM technique: what is the biomarker of each disorder? BMC Neurosci. 2022;23(1):48. doi: 10.1186/s12868-022-00725-9.

38. Bandt SK, de Rochefort L, Chen WW, Dimov AV, Spincemaille P, Kopell BH, et al. Clinical Integration of Quantitative Susceptibility Mapping Magnetic Resonance Imaging into Neurosurgical Practice. World Neurosurg. 2019;122:E10–E9. doi: 10.1016/j.wneu.2018.08.213.

39. Agnello F, Albano D, Micci G, Di Buono G, Agrusa A, Salvaggio G, et al. CT and MR imaging of cystic renal lesions. Insights Imaging. 2020;11(1):5. doi: 10.1186/s13244-019-0826-3.

40. Balci NC, Semelka RC, Patt RH, Dubois D, Freeman JA, Gomez-Caminero A, Woosley JT. Complex renal cysts: findings on MR imaging. AJR Am J Roentgenol. 1999;172(6):1495–500. doi: 10.2214/ajr.172.6.10350279.

41. Bae KT, Grantham JJ. Imaging for the prognosis of autosomal dominant polycystic kidney disease. Nat Rev Nephrol. 2010;6(2):96–106. doi: 10.1038/nrneph.2009.214.

42. Sallee M, Rafat C, Zahar JR, Paulmier B, Grunfeld JP, Knebelmann B, Fakhouri F. Cyst infections in patients with autosomal dominant polycystic kidney disease. Clin J Am Soc Nephrol. 2009;4(7):1183–9. doi: 10.2215/CJN.01870309.

43. Gibson P, Watson ML. Cyst infection in polycystic kidney disease: a clinical challenge. Nephrol Dial Transplant. 1998;13(10):2455–7. doi: 10.1093/ndt/13.10.2455.

44. Carpenter JP, He T, Kirk P, Roughton M, Anderson LJ, de Noronha SV, et al. Calibration of myocardial T2 and T1 against iron concentration. J Cardiovasc Magn Reson. 2014;16(1):62. doi: 10.1186/s12968-014-0062-4.

45. Kamman RL, Go KG, Brouwer W, Berendsen HJ. Nuclear magnetic resonance relaxation in experimental brain edema: effects of water concentration, protein concentration, and temperature. Magn Reson Med. 1988;6(3):265–74. doi: 10.1002/mrm.1910060304.

46. Jensen JH, Chandra R. Strong field behavior of the NMR signal from magnetically heterogeneous tissues. Magn Reson Med. 2000;43(2):226–36. doi: 10.1002/(sici)1522-2594(200002)43:2<226::aid-mrm9>3.0.co;2-p.

47. Luz Z, Meiboom S. Nuclear Magnetic Resonance Study of Protolysis of Trimethylammonium Ion in Aqueous Solution - Order of Reaction with Respect to Solvent. J Chem Phys. 1963;39(2):366-&. doi: Doi 10.1063/1.1734254.

48. Frahm J, Haase A, Matthaei D. Rapid NMR imaging of dynamic processes using the FLASH technique. Magn Reson Med. 1986;3(2):321–7. doi: 10.1002/mrm.1910030217.

49. Bernstein MA, King KF, Zhou ZJ. Handbook of MRI pulse sequences. Amsterdam ; Boston: Academic Press; 2004.

50. Shi H, Jia J, Li D, Wei L, Shang W, Zheng Z. Blood oxygen level-dependent magnetic resonance imaging for detecting pathological patterns in patients with lupus nephritis: a preliminary study using gray-level co-occurrence matrix analysis. J Int Med Res. 2018;46(1):204–18. doi: 10.1177/0300060517721794.

51. Mie MB, Nissen JC, Zollner FG, Heilmann M, Schoenberg SO, Michaely HJ, Schad LR. Susceptibility weighted imaging (SWI) of the kidney at 3T--initial results. Z Med Phys. 2010;20(2):143–50. doi: 10.1016/j.zemedi.2010.02.004.

52. Pan L, Chen J, Xing W, Xing Z, Zhang J, Peng Y, Zhang Z. Magnetic resonance imaging evaluation of renal ischaemia-reperfusion injury in a rabbit model. Exp Physiol. 2017;102(8):1000–6. doi: 10.1113/EP086203.

53. Zhang JG, Xing ZY, Zha TT, Tian XJ, D. YN, Chen J, Xing W. Longitudinal assessment of rabbit renal fibrosis induced by unilateral ureteral obstruction using two-dimensional susceptibility weighted imaging. J Magn Reson Imaging. 2018;47(6):1572–7. doi: 10.1002/jmri.25915.

54. Li J, Chang S, Liu T, Wang Q, Cui D, Chen X, et al. Reducing the object orientation dependence of susceptibility effects in gradient echo MRI through quantitative susceptibility mapping. Magn Reson Med. 2012;68(5):1563–9. doi: 10.1002/mrm.24135.

55. Yablonskiy DA, Haacke EM. Theory of NMR signal behavior in magnetically inhomogeneous tissues: the static dephasing regime. Magn Reson Med. 1994;32(6):749–63. doi: 10.1002/mrm.1910320610.

56. Dimov AV, Li J, Nguyen TD, Roberts AG, Spincemaille P, Straub S, et al. QSM Throughout the Body. J Magn Reson Imaging. 2023. doi: 10.1002/jmri.28624.

57. White RM. Quantum theory of magnetism : magnetic properties of materials. 3rd, completely rev. ed. Springer series in solid-state sciences,. Berlin ; New York: Springer; 2007.

58. Pauling L. General chemistry. New York: Dover Publications, Inc.; 1988.

59. Kazancioglu R, Ecder T, Altintepe L, Altiparmak MR, Tuglular S, Uyanik A, et al. Demographic and clinical characteristics of patients with autosomal dominant polycystic kidney disease: a multicenter experience. Nephron Clin Pract. 2011;117(3):c270–5. doi: 10.1159/000320745.

60. Nishiura JL, Neves RF, Eloi SR, Cintra SM, Ajzen SA, Heilberg IP. Evaluation of nephrolithiasis in autosomal dominant polycystic kidney disease patients. Clin J Am Soc Nephrol. 2009;4(4):838–44. doi: 10.2215/CJN.03100608.

61. Torres VE, Wilson DM, Hattery RR, Segura JW. Renal stone disease in autosomal dominant polycystic kidney disease. Am J Kidney Dis. 1993;22(4):513–9. doi: 10.1016/s0272-6386(12)80922-x.

62. Yaman S, Tekin HC. Magnetic Susceptibility-Based Protein Detection Using Magnetic Levitation. Anal Chem. 2020;92(18):12556–63. doi: 10.1021/acs.analchem.0c02479.

63. Chang S, Zhang J, Liu T, Tsiouris AJ, Shou J, Nguyen T, et al. Quantitative Susceptibility Mapping of Intracerebral Hemorrhages at Various Stages. J Magn Reson Imaging. 2016;44(2):420–5. doi: 10.1002/jmri.25143.

64. Bradley WG, Jr. MR appearance of hemorrhage in the brain. Radiology. 1993;189(1):15–26. doi: 10.1148/radiology.189.1.8372185.

65. Langkammer C, Liu T, Khalil M, Enzinger C, Jehna M, Fuchs S, et al. Quantitative susceptibility mapping in multiple sclerosis. Radiology. 2013;267(2):551–9. doi: 10.1148/radiol.12120707.

66. Choi Y, Jang J, Kim J, Nam Y, Shin NY, Ahn KJ, et al. MRI and Quantitative Magnetic Susceptibility Maps of the Brain after Serial Administration of Gadobutrol: A Longitudinal Follow-up Study. Radiology. 2020;297(1):143–50. doi: 10.1148/radiol.2020192579.

67. Liu S, Wang C, Zhang X, Zuo P, Hu J, Haacke EM, Ni H. Quantification of liver iron concentration using the apparent susceptibility of hepatic vessels. Quant Imaging Med Surg. 2018;8(2):123–34. doi: 10.21037/qims.2018.03.02.

68. Wei H, Dibb R, Zhou Y, Sun Y, Xu J, Wang N, Liu C. Streaking artifact reduction for quantitative susceptibility mapping of sources with large dynamic range. NMR Biomed. 2015;28(10):1294–303. doi: 10.1002/nbm.3383.

69. Liu Z, Kee Y, Zhou D, Wang Y, Spincemaille P. Preconditioned total field inversion (TFI) method for quantitative susceptibility mapping. Magn Reson Med. 2017;78(1):303–15. doi: 10.1002/mrm.26331.

70. Kim W, Shin HG, Lee H, Park D, Kang J, Nam Y, et al. chi-Separation Imaging for Diagnosis of Multiple Sclerosis versus Neuromyelitis Optica Spectrum Disorder. Radiology. 2023;307(1):e220941. doi: 10.1148/radiol.220941.

71. Chen J, Gong NJ, Chaim KT, Otaduy MCG, Liu C. Decompose quantitative susceptibility mapping (QSM) to sub-voxel diamagnetic and paramagnetic components based on gradient-echo MRI data. Neuroimage. 2021;242:118477. doi: 10.1016/j.neuroimage.2021.118477.

72. Dimov AV, Nguyen TD, Gillen KM, Marcille M, Spincemaille P, Pitt D, et al. Susceptibility source separation from gradient echo data using magnitude decay modeling. J Neuroimaging. 2022;32(5):852–9. doi: 10.1111/jon.13014.

73. Irazabal MV, Rangel LJ, Bergstralh EJ, Osborn SL, Harmon AJ, Sundsbak JL, et al. Imaging classification of autosomal dominant polycystic kidney disease: a simple model for selecting patients for clinical trials. J Am Soc Nephrol. 2015;26(1):160–72. doi: 10.1681/ASN.2013101138.

