## Supplementary figures and images for "Quantitative susceptibility mapping for detection of kidney stones, hemorrhage differentiation and cyst classification in ADPKD"

### Supplemental figure S1

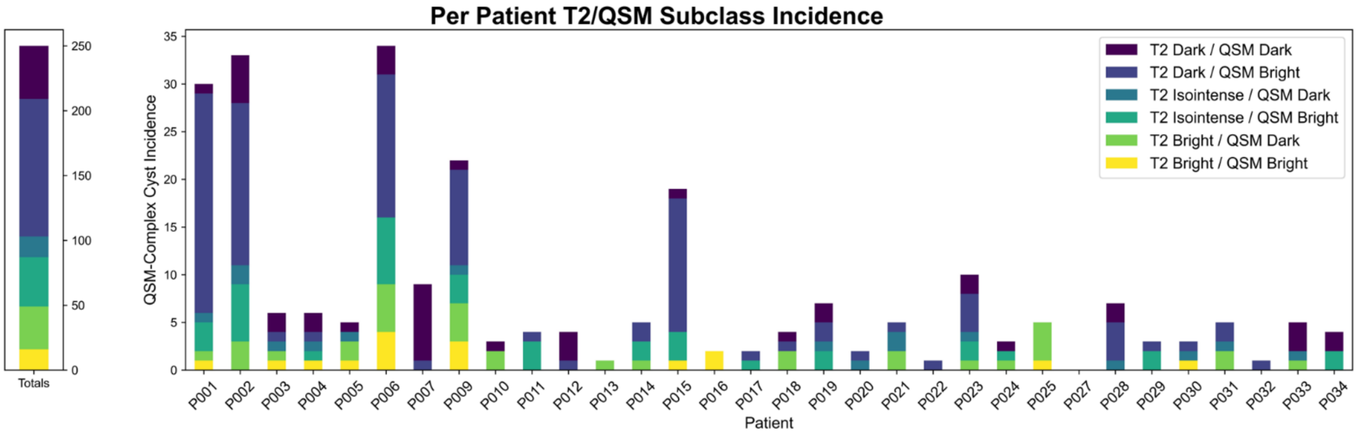


Figure S1. Distribution of the T2/QSM cyst subtypes across individual subjects
