## Supplemental figure S2 for "Quantitative susceptibility mapping for detection of kidney stones, hemorrhage differentiation and cyst classification in ADPKD"

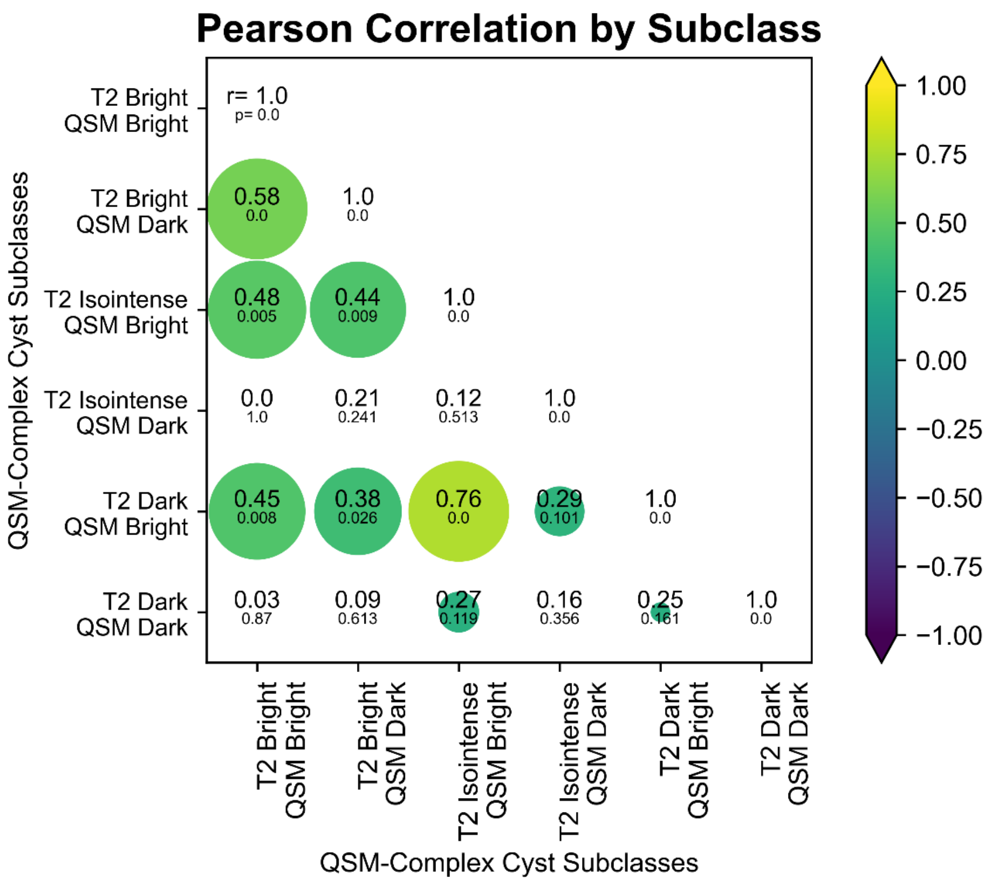


Figure S2. Pearson correlation matrix demonstrating linear correlation between the incidence of each cyst subclass across all patients. The color of the circles indicates magnitude of the correlation coefficient, while their size represents its statistical significance.
